# Peripheral inflammatory response in human tuberculosis treatment is predicted by a combination of pathogen sterilization and microbiome dysbiosis

**DOI:** 10.1101/2020.02.25.20027870

**Authors:** Matthew F. Wipperman, Shakti K. Bhattarai, Charles Kyriakos Vorkas, Ying Taur, Laurent Mathurin, Katherine McAulay, Stalz Charles Vilbrun, Daphie Jean Francois, James Bean, Kathleen F. Walsh, Carl Nathan, Daniel W. Fitzgerald, Michael S. Glickman, Vanni Bucci

## Abstract

Antibiotic therapy cures infection predominantly by killing the infecting pathogen, but for infections such as tuberculosis (TB), which are accompanied by chronic inflammation, the salutary effects of antibiotic therapy may reflect a combination of pathogen killing and microbiome alteration. This question has not been examined in humans due to the difficulty in dissociating the immunologic effects of antibiotic induced pathogen clearance and microbiome alteration. We analyzed sputum TB bacterial load, microbiome composition, and peripheral blood transcriptomics from a clinical trial (NCT02684240) comparing two antimicrobial therapies for tuberculosis, only one of which was clinically effective. We confirm that standard TB therapy (HRZE) rapidly depletes Clostridia from the intestinal microbiota. The antiparasitic drug nitazoxanide (NTZ), although ineffective in reducing *Mycobacterium tuberculosis* (*Mtb*) bacterial load in the sputum, caused profound alterations to host microbiome composition overlapping with alterations generated by HRZE. We then evaluated the effect of these two treatments on the TB driven inflammatory state and found that whereas HRZE normalized proinflammatory TB-associated gene sets, NTZ exacerbated these pathways. Using Random Forest Regression, we identify both pathogen sterilization and microbiome disruption as the top predictors of changes in TB-associated inflammatory transcriptomic markers. We then validate the observed microbiome-peripheral gene expression associations in an independent human cohort of healthy subjects in which the abundance of Clostridia was positively associated with homeostatic, and negatively associated with pro-inflammatory pathways, while the abundance of Bacilli and Proteobacteria species displayed the opposite trend. Our findings indicate that antibiotic-induced reduction in pathogen burden and changes in the microbiome are independently associated with treatment-induced changes of the inflammatory response of active TB, and more broadly indicate that response to antibiotic therapy may be a combined effect of pathogen killing and microbiome driven immunomodulation. Additionally, to our knowledge, this is the first analysis to directly test the hypothesis that the microbiome composition is associated with peripheral gene expression inflammatory profile in humans.

## Introduction

There is mounting evidence that the gut microbiome has an important role in the modulation of host physiology, with a wealth of studies having associated microbiome composition and functions with differential inflammatory, neurological, and even behavioral activity^1^. Gastrointestinal colonization by specific taxa with particular metabolic capacities has been shown to differentially modulate host biology^2^. For example, colonization by a subset of Clostridia enhanced anti-inflammatory phenotypes in mice^3^, and enrichment in specific members of the *Bacteroides* and *Parabacteroides* genera induced CD8+ T cell responses and anticancer activity in mice and marmosets^4^, as well as correlating with the abundance of these immune effectors in humans^5^. A multitude of experiments in mice have allowed for the determination of mechanisms by which intestinal mucosal-associated bacteria affect host physiology at the epithelial interface and systemically throughout their host ^6,7^.

Despite these observations, it is unknown whether, and to what degree, microbiome changes are responsible for changes in human systemic inflammatory responses. This knowledge gap is due in part to the difficulty of isolating the microbiome dependent effects from other aspects of human physiology and in discerning the direction of causality in human studies. As microbial communities in the gut promote the development and maintenance of innate and adaptive immune responses, including microbiota-educated immune cells and many small molecules that circulate throughout the periphery, we would expect to observe both localized and systemic host effects due to microbiome alterations ^8^.

Infection by *Mycobacterium tuberculosis* (*Mtb*) is the cause of tuberculosis (TB) disease—the 9^th^ leading cause of death on Earth^9^. A plethora of studies using whole blood transcriptomics have documented that individuals with active TB display a different systemic gene expression pattern compared to people with latent disease, other respiratory diseases, or no known infection^9-11^. Specifically, infection with *Mtb* leads to heightened expression of inflammatory pathways, most notably the Type I and Type II interferon pathways^12-15^, and this pattern resolves with antibiotic therapy^12,15,16^. A recent meta-analysis combining microarray and RNAseq data from studies aimed at identify active TB transcriptional signatures confirmed the findings about a specific set of peripheral blood transcripts that are biomarkers of active TB disease, relative to healthy individual, or those with latent TB infection (LTBI)^17^.

Antibiotic treatment for active TB involves combination therapy with narrow spectrum and prodrug agents with mostly *Mycobacterial*-specific targets. The World Health Organization guidelines for treating infection with drug sensitive *Mtb* are to give isoniazid (H), rifampin (R), pyrazinamide (Z), and ethambutol (E) (HRZE) for two months and then to continue HR for an additional four months. The effects of HRZE therapy on the intestinal microbiome were demonstrated in a longitudinal study in mice^18^ and cross-sectional study in humans^19^, which indicated that the major phyla perturbed are from the class Clostridia, a group of obligate anaerobes in the gut with well described immunomodulatory effects on the host^2,3,20,21^. Given that HRZE treatment causes microbiome shifts that include the depletion of many Clostridia species, and given the role that these species play in modulation of host biology in mice and humans, we reasoned that there could be a connection between the microbiome alterations observed during HRZE therapy and the resolution of systemic inflammatory responses to TB. However, because HRZE therapy rapidly reduces the burden of *Mtb* in the early phase of treatment, it is difficult to uncouple the immunologic effects of pathogen killing from microbiome perturbation without a control group that has either pathogen killing or microbiome perturbation, but not both.

An opportunity to address this issue arose when we analyzed secondary endpoint data from a clinical trial (NCT02684240) that compared the early bactericidal effect (EBA) of standard TB therapy with HRZE to the antiparasitic drug nitazoxanide (NTZ), recently shown to possess antimycobacterial activity *in vitro*^22,23^. We found that NTZ perturbed the intestinal microbiome, with pathobiont domination and Clostridia depletion. This contrasted with HRZE treatment, which had a narrow effect on intestinal Clostridia after only two weeks of treatment. We then found that HRZE and NTZ had distinct effects on host peripheral gene expression, with HRZE resolving interferon signatures and NTZ exacerbating them. We used machine learning to determine the factors that predict correction or exacerbation of TB associated systemic inflammation and found that the three most important predictors are (a) the *Mtb* level in the sputum, (b) the abundance of Clostridia species that associate with inflammatory renormalization, and (c) the abundance of antibiotic-promoted Proteobacteria that associate with exacerbation of proinflammatory pathways. We next investigated these relationships using a validation cohort of healthy Haitian community controls and household contacts of TB patients, previously described^5^. Using machine learning to investigate relationships between the microbiome and peripheral gene pathways derived from the MiSigDB hallmark gene pathways database, we validated many of the relationships between peripheral gene expression and microbiome composition. We believe these results provide support to the oft stated hypothesis that there exists clear regulatory relationships between gut microbiome composition and peripheral composition, at both the immune ^5^, and gene regulatory levels in humans.

## Results

### Gut microbiome diversity is depleted after two weeks of HRZE or NTZ treatment

As detailed elsewhere, the GHESKIO centers in Port au Prince, Haiti conducted a prospective, randomized, early bactericidal activity (EBA) study in treatment-naive, drug-susceptible adult patients with uncomplicated pulmonary tuberculosis (TB) (ClinicalTrials.gov Identifier: NCT02684240)^23^. Participants were randomized to receive either NTZ, 1000 mg po (oral) twice daily, or standard oral therapy with isoniazid 300 mg daily, rifampin 600 mg daily, pyrazinamide 25 mg/kg daily, and ethambutol 15 mg/kg daily (referred to as HRZE) for 14 days (Figure 1A). The primary endpoint of the trial was sputum bacterial load (measured by time to culture positivity, TTP) in a BACTEC liquid culture system. Sputum was collected from 6pm to 9am every other day to quantify mycobactericidal activity of each treatment regimen. As reported^23^, HRZE resulted in a predictable increase in the TTP (corresponding to reduced bacterial load) over the first two weeks of therapy compared to pretreatment TTP, consistent with its known potent bactericidal activity. However, NTZ, despite potent in vitro activity^24^, did not have any significant effect on TTP after 14 days (Figure 1B)^23^. This lack of NTZ efficacy was traced to a failure of the drug to penetrate the sputum^23^. All patients were subsequently switched to HRZE standard treatment.

**Figure 1:**
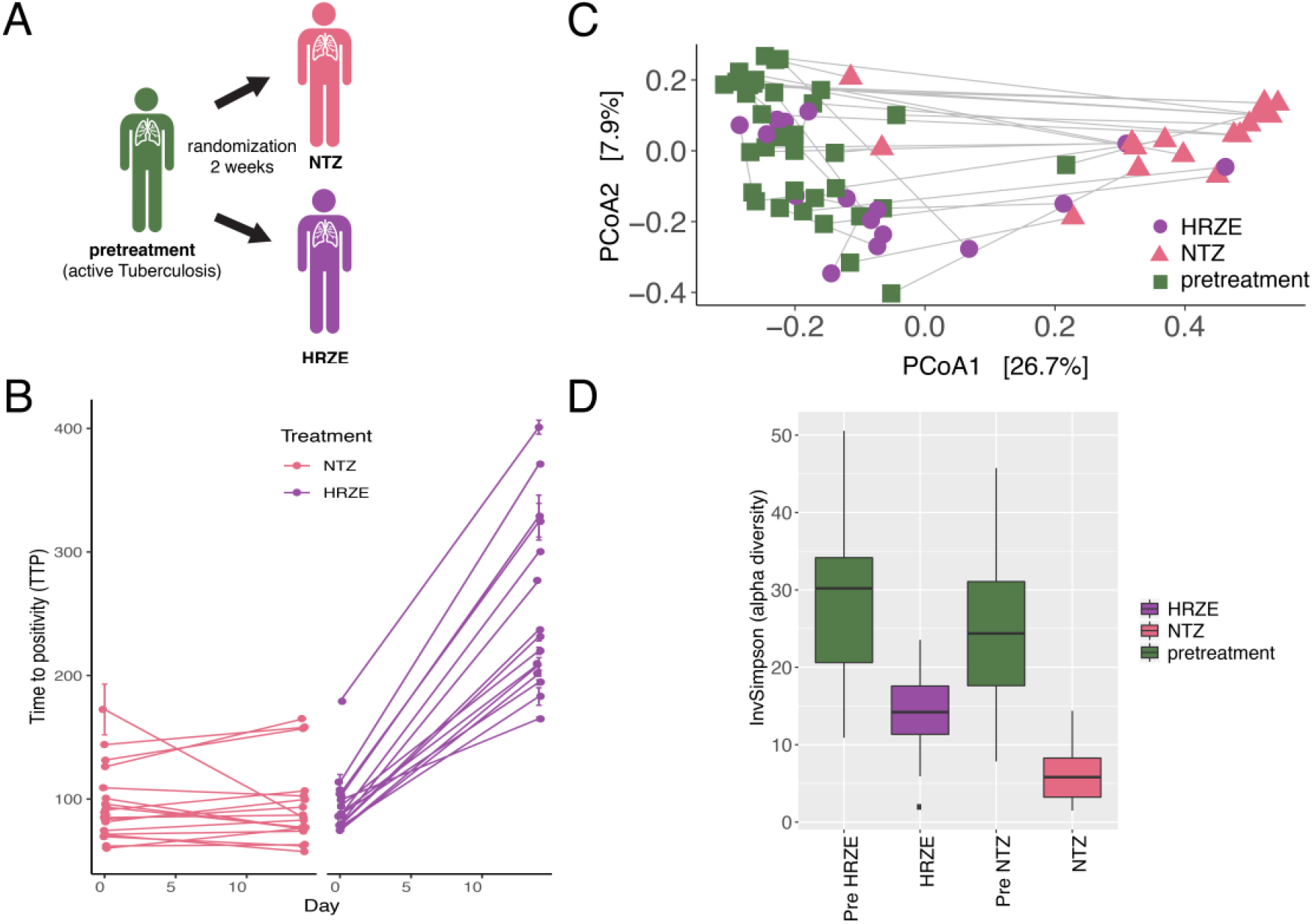
Both HRZE and NTZ perturb the gut microbiome after two weeks of therapy. **A**. Schematic showing active TB patients randomized to either HRZE (standard of care) or NTZ. **B**. Paired time to positivity (TTP) at day 0 and day 14 for the NTZ treatment cohort and HRZE treatment cohort. **C**. Principal components analysis (PCoA) with Bray-Curtis distance showing differences in microbiome community structure between individuals before and after 14 days of either HRZE or NTZ treatment. The grey line connects pre and day 14 treatment paired samples. **D**. Microbiota alpha diversity plotted using the Inverse Simpson index. There was no significant difference between the pretreatment groups, and both groups had significantly (p < 0.01, Wilcoxon signed-rank test) reduced alpha diversity after 14 days of treatment.

We have reported^19^ that HRZE therapy depletes members of the order Clostridiales, but the cross-sectional design of that study did not allow for conclusions about the rapidity of this effect and most importantly, did not include pretreatment samples that will allow for assessment of baseline microbiome composition. To investigate microbiome changes induced by NTZ or HRZE, we extracted and amplified bacterial and archaeal DNA using V4 – V5 16S rDNA sequencing. Stool samples were collected before the start of treatment and on day 14 of therapy (Figure 1A). Using principal components analysis (PCoA) with Bray Curtis distances, we determined that antibiotic administration induced rapid changes in microbiome community structure after two weeks in both the NTZ and HRZE groups (PERMANOVA: p<0.001 and p<0.02, respectively), compared with pretreatment (Figure 1C). HRZE samples clustered closer to pretreatment samples than did NTZ to pretreatment. Both treatments were characterized by a significant drop in alpha diversity (the Inverse Simpson index) when compared to pretreatment samples, and despite expected interindividual heterogeneity, NTZ treatment samples had a significantly lower diversity compared to HRZE (Figure 1C). There was no significant difference between the day 0 diversity metrics between the pre-randomized active TB groups (Figure 1D).

### Overlapping taxonomic alterations in microbiome composition induced by NTZ and HRZE

Taxonomic profiling of differential amplicon sequence variants (ASVs) obtained from the 16S rDNA profiling between the pretreatment and antibiotic groups revealed that, whereas all trial participants began with typical heterogeneous community structure, after two weeks of treatment with HRZE, members of the class Clostridia were depleted, leaving most other clades unaltered (Figure 2A,B). In contrast, NTZ had a much more pronounced and broad effect compared to HRZE (Figure 2A,C). Specifically, NTZ not only reduced the relative abundance of a greater number of Clostridia compared to HRZE, but also led to the increase of aerobic and facultative anaerobic pathobiont organisms such as *Escherichia* and *Klebsiella* (Proteobacteria). Additionally, several participants became dominated (with domination defined as >30% relative abundance according to Taur, et al^25^) by single clades of *Actinobacteria* or *Bacilli* in the NTZ arm of the trial (Figure 2A,C and Supplementary Figure S1 and S2).

**Figure 2:**
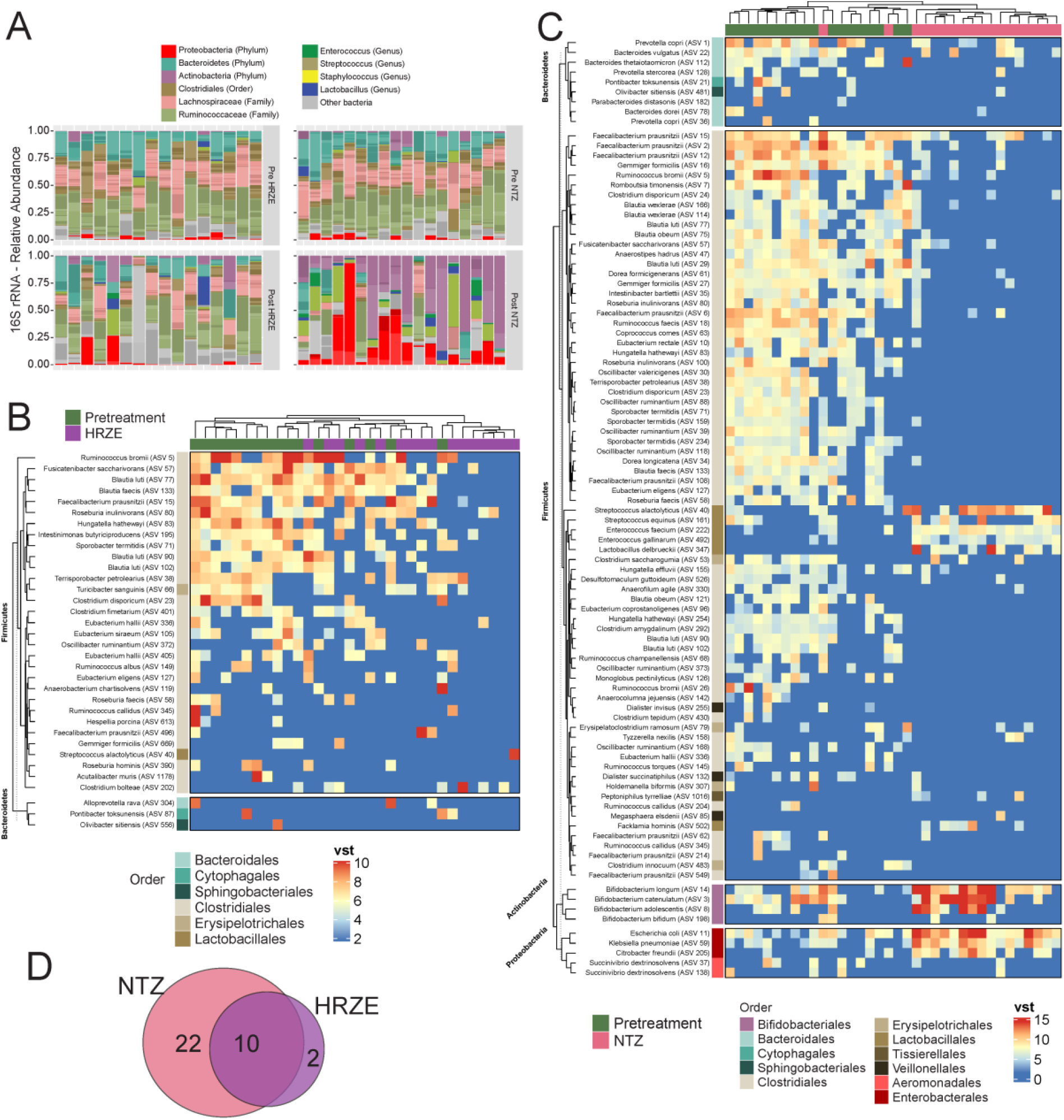
Overlapping and distinct microbiome perturbation induced by NTZ and HRZE. **A**. Relative abundance showing microbiota composition in each individual patient as stacked paired samples corresponding to pre and post treatment. Only paired samples were used for these analyses. **B**. Unsupervised hierarchical clustering of the abundances of ASVs identified to be significantly affected by HRZE treatment (p<0.01); VST indicates variance stabilized transformed counts from DESeq2. **C**. Unsupervised hierarchical clustering of the abundances of ASVs identified to be significantly affected by NTZ treatment (p<0.01). **D**. Number of Clostridia genera depleted (i.e. reduced in relative abundance) by HRZE and NTZ treatment.

Comparison of the overlap of the two antibiotic perturbations with respect to number of Clostridia genera affected (i.e., testing for differences pre-post HRZE/NTZ administration on bacterial abundances grouped at the genus level) revealed that 83% (10/12) of the HRZE-depleted Clostridia were also depleted by NTZ (Figure 2D). HRZE uniquely affected Clostridia genera *Acutalibacter* and *Hespellia*, whereas NTZ affected 22 unique Clostridia genera, including *Blautia, Dorea, Eubacterium, Faecalibacterium, Oscillibacter, Ruminococcus*, and *Lachnospiraceae*. Both treatments affected the genera *Clostridium, Fusicatenibacter, Hungatella, Intestinibacter, Intestinimonas, Kineothrix, Roseburia, Sporobacter, Terrisporobacter*, and an uncultured member of the Family *Ruminococcaceae*. Taken together, these data demonstrate that NTZ had a more severe disruptive effect on the intestinal microbiota than HRZE and that most of the HRZE effects on the microbiota (e.g. loss of Class Clostridia) were also evident in NTZ treated subjects.

### HRZE and NTZ uniquely affect host peripheral gene expression

To determine how inflammatory transcriptional signatures in peripheral blood are affected by TB treatment, we applied unsupervised (PCA) and supervised learning (Support Vector Machine and Random Forest Classification) methods to the DESeq-normalized RNA transcript abundances from RNA derived from peripheral blood (Supplementary Figure S3). This analysis showed that, overall, HRZE treatment caused a more substantial change (in terms of the number of genes affected) in expression profile than NTZ when comparing pretreatment to 2 week treatment transcriptome samples, as measured by random forest/support vector machine ROC curves or principal component analysis (Supplementary Figure S3A-B).

To identify specific RNA transcripts affected by each treatment, we used DESeq analysis on the paired-sample transcript abundance of RNA and compared pretreatment samples to samples obtained at day 14 after HRZE (n=8) or NTZ treatment (n=14) independently. The paired nature of samples was considered in the analysis to account for person-specific baseline normalization. We found that 2 weeks of HRZE treatment was associated with changes in 1374 transcripts at a p-value cutoff of p<0.05 and 503 at a p-value cutoff of p<0.001 (throughout, all p-values from DESeq are adjusted using the Benjamini-Hochberg method for multiple comparisons). Repeating the same analysis for NTZ-treated individuals, we identified 811 differentially expressed genes at a p-value cutoff of p<0.05 and only 15 at p<0.001.

We determined the functional pattern of treatment-induced changes in overall transcript abundance, by performing gene set enrichment analysis (GSEA)^26^ (See Methods) using the ranked DESeq gene expression data for the differentially expressed genes in each arm separately. In the HRZE arm we observed reduced expression of the pathways of inflammatory response, IFNγ response, IFNα response, TNFα signaling via NFκB, and IL6 JAK STAT3, all of which are consistent with the immunologic effects of antibiotic induced reduction in the levels of the bacterial pathogen, given the demonstrated relevance of these signaling pathways to pathogenesis^12,15,27^ (Figure 3A). In contrast, NTZ treatment, which perturbed the microbiome without significantly affecting *Mtb* bacterial load in the sputum, had the opposite effect. Inflammatory signaling pathways reduced by HRZE, including TNFα signaling, IFNγ signaling, and type 1 interferon signaling, were all enriched by NTZ treatment (Figure 3A). Several other pathways such as hypoxia, apoptosis, and reactive oxygen species (ROS), which are considered hallmarks of immune dysregulation^28^, were also enriched by NTZ treatment (Figure 3B). When comparing the list of transcripts significantly affected by either HRZE (1374) or NTZ (811), after taking into account person-specific gene normalization, we found that only 86 genes were affected by both treatments (p < 0.05). Eighty of these genes had the same pre/post treatment pattern in HRZE and NTZ-treated individuals, thus leaving 1294 (94.2%) and 731 (90.1%) to be HRZE and NTZ-specific changes to peripheral gene expression (Figure 3C). Of the 86 genes affected by both HRZE and NTZ, only 10 were reported to be peripheral blood transcriptomic markers of active tuberculosis ^12,13,15^. Interestingly, six active-TB associated genes belong to the group of 80 genes that similarly respond to HRZE and NTZ (7.5%). The remaining four TB-related genes (P2RY14, ADM, CARD16, DHRS9) belong to the group of six genes (P2RY14, NRN1, ADM, JAG1, CARD16, DHRS9) that were reduced in expression and renormalized with HRZE but increased and exacerbated TB signature with NTZ (67%) (Figure 3C).

**Figure 3:**
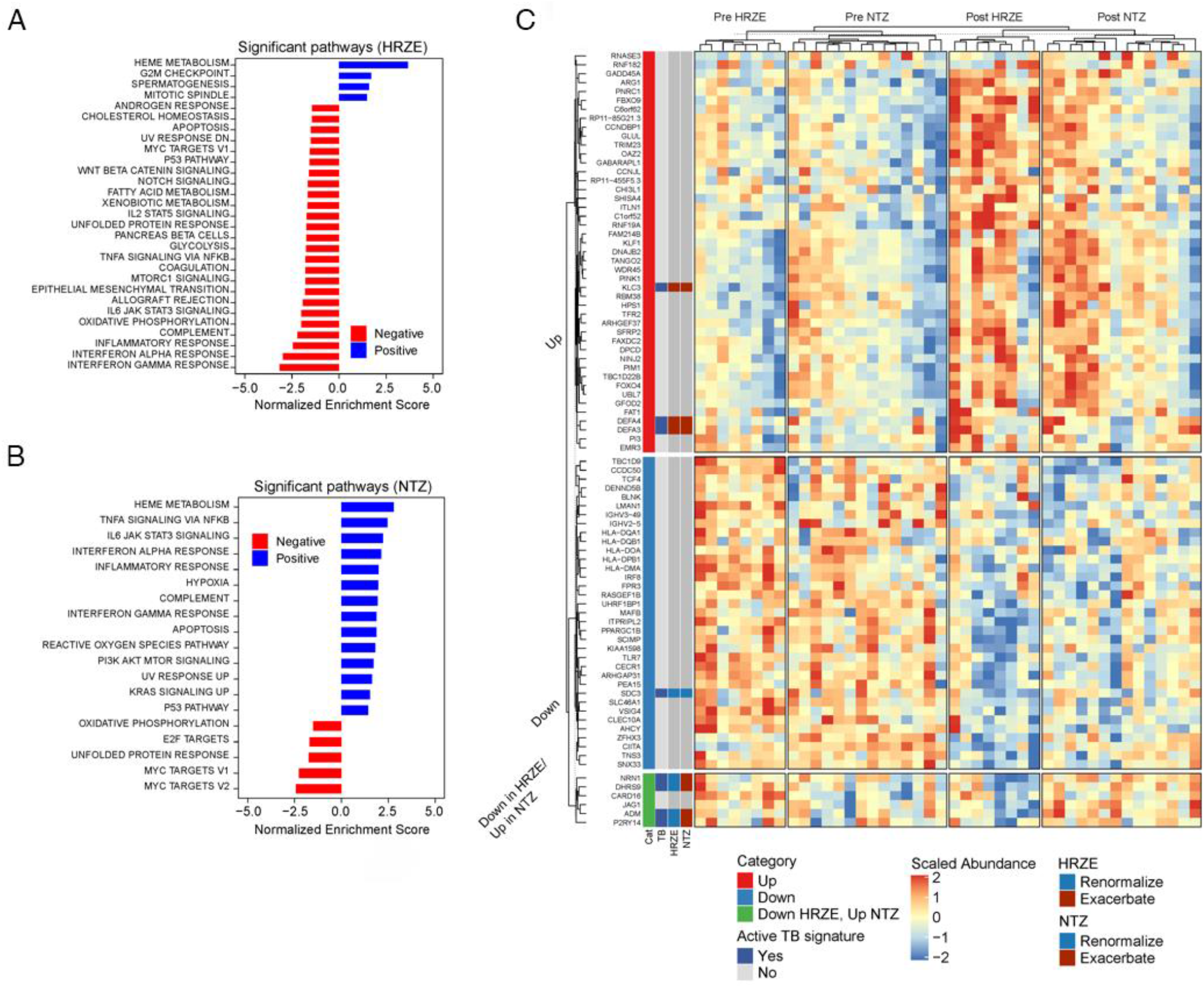
Hallmark pathway gene set enrichment analysis and gene expression comparison in HRZE and NTZ treated cohorts. **A**,**B**. Hallmark gene pathway changes associated with 2 weeks of HRZE (A) or NTZ (B). Positive are pathways overrepresented at 2 weeks of therapy, and negative are pathways underrepresented at 2 weeks, both compared to pretreatment. All pathways are significant (p<0.05) with the associated Normalized Enrichment Score (NES) shown on the × axis, which considers pathway size. **C**. Overlap in genes that are differentially altered in both HRZE and NTZ treatments compared to pretreatment. Transcripts annotated in the left column in red (Up) rise with HRZE and NTZ. Transcripts annotated in blue (Down) are suppressed by both HRZE and NTZ. Genes annotated in green are down in HRZE and up in NTZ. The second column of vertical metadata scores each gene as either present or absent in prior active TB transcriptional signatures.

To focus our analysis on validated transcriptomic markers of active TB from prior studies, we examined the recently reported list of 373 transcripts that have been associated and validated in multiple human cohorts on multiple sequencing platforms (microarray and RNAseq) to be differentially abundant between active and healthy control and/or active and LTBI individuals^17^ in a comprehensive meta-analysis. In our study, we detected 361 of these 373 transcripts in pretreatment active TB subjects. We defined three classes of changes to these transcripts with two weeks of HRZE or NTZ treatment: 1) *renormalization* (transcripts whose pre-post HRZE/NTZ foldchange in expression displays the same sign (or direction) of the previously-reported fold-change between active TB and control/LTBI; 2) *unchanged* (transcripts with no change in expression between pre-post HRZE/NTZ administration); and 3) *exacerbation* (genes whose pre-post HRZE/NTZ fold-change sign is opposite to the previously-reported fold-change between active TB and control/LTBI). Of the 361 transcripts, 173 (48%) were found to be affected by HRZE. Of these 173, 151 (87%) renormalize with HRZE treatment, whereas 22 (13%) exacerbate (Supplementary Table S1, Figure 4). In contrast, NTZ was found not only to have a smaller overall effect on the active TB signature, but also to only contribute to exacerbation. Specifically, only 28 genes were affected by NTZ (8%), of which 26 (96%) were in the exacerbation category (Supplementary Table S1, Figure 4).

**Figure 4:**
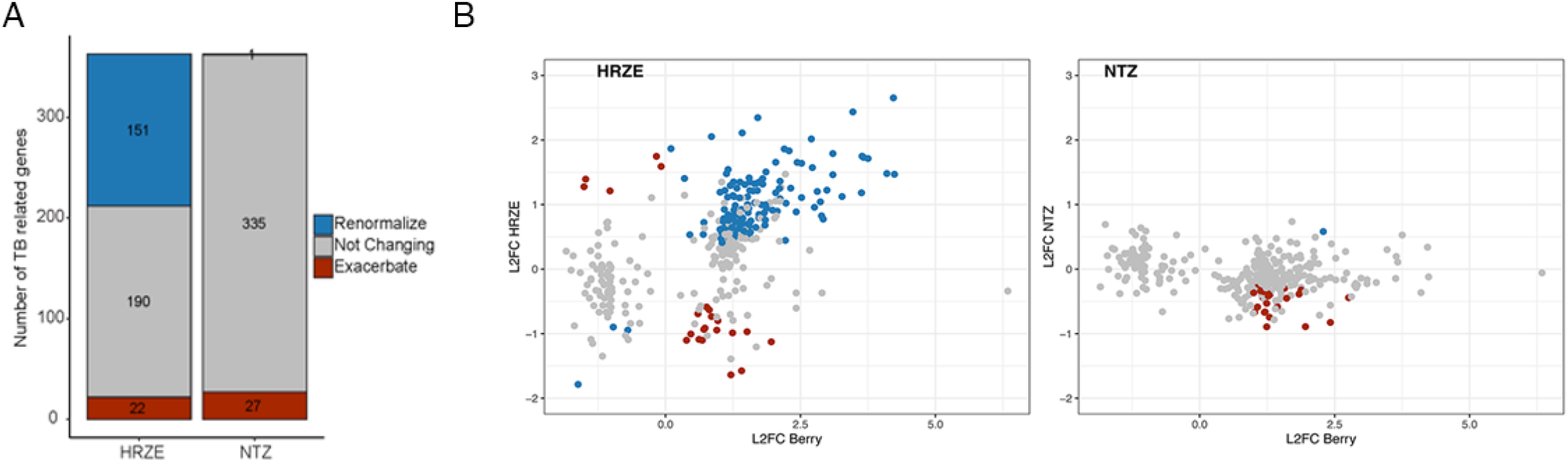
Effect of HRZE and NTZ on active TB transcriptional signatures. **A**. Summary of the number of active TB genes that renormalize (blue), do not change (grey), or exacerbate (red) as a result of each drug administration. **B**. Comparison of log2 Fold Change in gene expression from^17^ where the 373 active TB signature was first introduced with the log2 Fold Change of the same genes pre-post HRZE (left) or NTZ (right) from this study. A significant positive correlation (Pearson’s p<0.01) is observed for the 151 genes that renormalize with HRZE treatment.

Taken together, our data suggest that both HRZE and NTZ lead to changes in the peripheral transcriptomic pattern of active TB, with HRZE having a more dramatic effect, at least as measured by the number of genes differentially expressed before and after two weeks of therapy compared to NTZ, both overall as well as when considering a previously-reported signature of active TB. Additionally, our analysis suggests that two weeks of HRZE treatment normalizes almost 50% of the peripheral gene expression profile characteristic of active tuberculosis, consistent with prior studies, whereas NTZ had the opposite effect: exacerbating the transcriptional pattern for a subset of transcripts that reflect disease activity.

### Relationships between gene expression and changes in the microbiome in the longitudinal treatment cohort

Resolution of disease manifestations by antibiotic therapy of a chronic, pro-inflammatory infection such as tuberculosis is likely to reflect a complex interplay of pathogen killing and resolution of inflammatory responses, which may occur with different temporal profiles^29^. Although a primary effect of antibiotics is killing of the pathogen, perturbation of the microbiome during therapy might also affect disease resolution due to an independent effect on peripheral inflammation, but these two effects are difficult to uncouple and therefore remain hypothetical. Because we observed changes in peripheral transcriptomic profiles by both HRZE and NTZ in active TB subjects, in some cases divergent between HRZE and NTZ, despite the fact that NTZ did not significantly reduce *Mtb* bacterial load in the sputum, this dataset provided a unique opportunity to investigate potential effects of antibiotic induced microbiome changes on disease driven inflammation. We hypothesized that TB therapy-induced perturbations of the gut microbiota that might be predictive of the observed changes in host gene expression.

To decouple the effect of microbiome perturbation from *Mtb*-killing in peripheral inflammatory transcriptomics, we used a machine learning approach. Specifically, we built random forest regression (RFR) models^30^ to predict the DESeq-normalized pre- and post-treatment expression profiles of each significantly differentially abundant transcript as a function of both *Mtb* bacterial load (TTP) (at day 0 and day 14) and of DESeq-normalized abundance of ASVs (microbiome components) (also at day 0 and day 14) found to be affected by each TB treatment, separately. We modeled data from each cohort independently because the treatment effects at the single gene and ASV level were distinct between each cohort and because at baseline there is no significant difference in microbiome and gene expression due to arm membership (PERMANOVA p > 0.05). We used the Boruta algorithm ^31^ for feature selection in predicting the expression profile of each gene (see Methods). We reasoned that this approach was appropriately suited for this type of “large p, small n” multi-omics dataset common in clinical research^32^. Several advantages of RFR modeling include: being agnostic to model structure (e.g. non-parametric regression), not having to meet common assumptions underlying classical regression techniques, and being able to intrinsically perform ranked feature selection. Importantly, while the interpretation of RFR is apparently less immediate compared to traditional regression (e.g. there are per-se no regression coefficients or betas), downstream analysis, which includes Permutated Importance ^33^ and accumulated local effects calculations^34^ (see Methods) allows for the estimation of the significance of predictors (e.g. TTP, microbiome constituents, etc.) as well as their effects on the dependent variable (e.g. host transcriptomic markers).

For both sets of HRZE- and NTZ-affected transcripts, we estimated the frequency of each feature to be identified as important by the RFR model. We performed this analysis considering all differentially expressed genes in both datasets (Supplementary Figure S4) as well as focusing only the 361 genes belonging to the list of 373 transcripts previously reported as an active TB signature. Remarkably, this analysis identified bacterial load and the abundance of members of Clostridia as the most important predictors of gene expression (Supplementary Figure S6A and 5B). For the HRZE arm (Supplementary Figure S6A), change in bacterial load (TTP) was the top predictor of the change in transcript abundance with treatment, which is found to be important in predicting the change in 75% of the 173 active TB transcripts and HRZE affected transcripts (Supplementary Figure S6). This is consistent with the idea that HRZE normalizes the active TB signature by reducing the pathogen burden in the lung (Supplementary Figure S6C). Surprisingly, even though sputum bacterial load did not change in aggregate in the NTZ cohort, TTP is still found to be the second most important feature in predicting changes in active TB gene expression, as it is found to contribute to the abundance of 48% of the 27 active TB and NTZ affected transcripts (Figure 3). We confirmed that the relationship between bacterial load and transcript abundance in the NTZ cohort (obtained via RFR) was a biological signal by performing linear-mixed effect modeling (lme in R) to predict the expression of each of these NTZ associated transcripts as a function of TTP and using patient ID as a random effect (Supplementary Figure S4) (model: lme(∼ TTP, random = ∼1|Patient.ID)). We found that despite the lack of clinical efficacy of the NTZ drug as measured by average TTP in the treated group, transcripts that are known to respond to TB bacterial load are altered in a direction that correlates with the TTP signal. This reflects the fact that while the magnitude of TTP change was not clinically significant across the entire NTZ cohort, person-specific changes in TTP were present in the NTZ cohort for TB-associated transcripts.

With respect to microbiome effects, Clostridia are predicted by both models to be the most important microbiome component predicting the change in the inflammatory transcriptomic pattern that reflects active TB and normalizes with treatment. Specifically, in the HRZE arm (Supplementary Figure S6A) *E. hallii* (ASV 336) and *S. termiditis* (ASV 71) are the second and third-best predictors of the abundance of over 50% of the HRZE-affected transcripts associated with active TB. In the NTZ arm (Supplementary Figure S6B) *G. formicilis (ASV 27)* is identified as the most important predictor with over 50% of NTZ-affected active TB transcripts predicted to co-vary with it. Of the other important microbiome features selected by the models, we observed the presence of a number of well characterized short chain fatty acid-producing Clostridia (including *F. prausnitizii, Clostrdium spp*., *B. lutii* in the HRZE group, or *D. longicatena* and *B. lutii* in the NTZ group), which are all depleted by the anti-TB treatment (Supplementary Figure S6C-D). Interestingly both models identified the same ASV, *Clostridium* XIVa *F. saccharivorans (ASV 57)* to be an important predictor of transcriptomic markers that are depressed by both treatments. Peculiar to the NTZ-data based model is the identification as important variables of *S. alactolyticus* (ASV 40), *E. faecium* (ASV 222) and *K. pneumoniae* (ASV 59) which are observed to significantly increased in abundance and dominate several of the NTZ-treated individuals (Figure 2).

Variable importance estimation techniques such as Boruta allow determination of the presence of significant associations among variables, but not their directionality (positive/negative effect). Therefore, we determined the direction of the contribution of each important feature to the abundance of each of the modeled active TB signature transcripts from the RFR modeling, by adapting the concept of accumulated local effects (ALE) calculations (see Methods)^34^ and using the sign of the average of the first-order derivative of the ALE plot as a binary indicator for direction. As most of these plots are monotonic and continuous curves, the first order derivative is a good approximation of the quantitative contribution of a hypothesized biologically relevant predictor to transcript abundance, in this case microbiota constituents or change in *Mtb* bacterial load (TTP). We displayed the sign of the *features x genes* matrices containing the first order derivative values (or slopes) as clustered heatmaps (Figure 5A-B). For both the HRZE and NTZ dataset-derived models, we observed that sputum bacterial load (TTP) clustered independently of the model-selected microbiome features (Figure 5A-B). The effect of reducing sputum bacterial load was predominantly to normalize gene expression in the relevant inflammatory pathways such as IFNγ and IFNα (negative contribution in heatmap for TTP means that rising TTP (i.e., lower bacterial load) is associated with normalizing transcripts) (Figure 5A).

**Figure 5:**
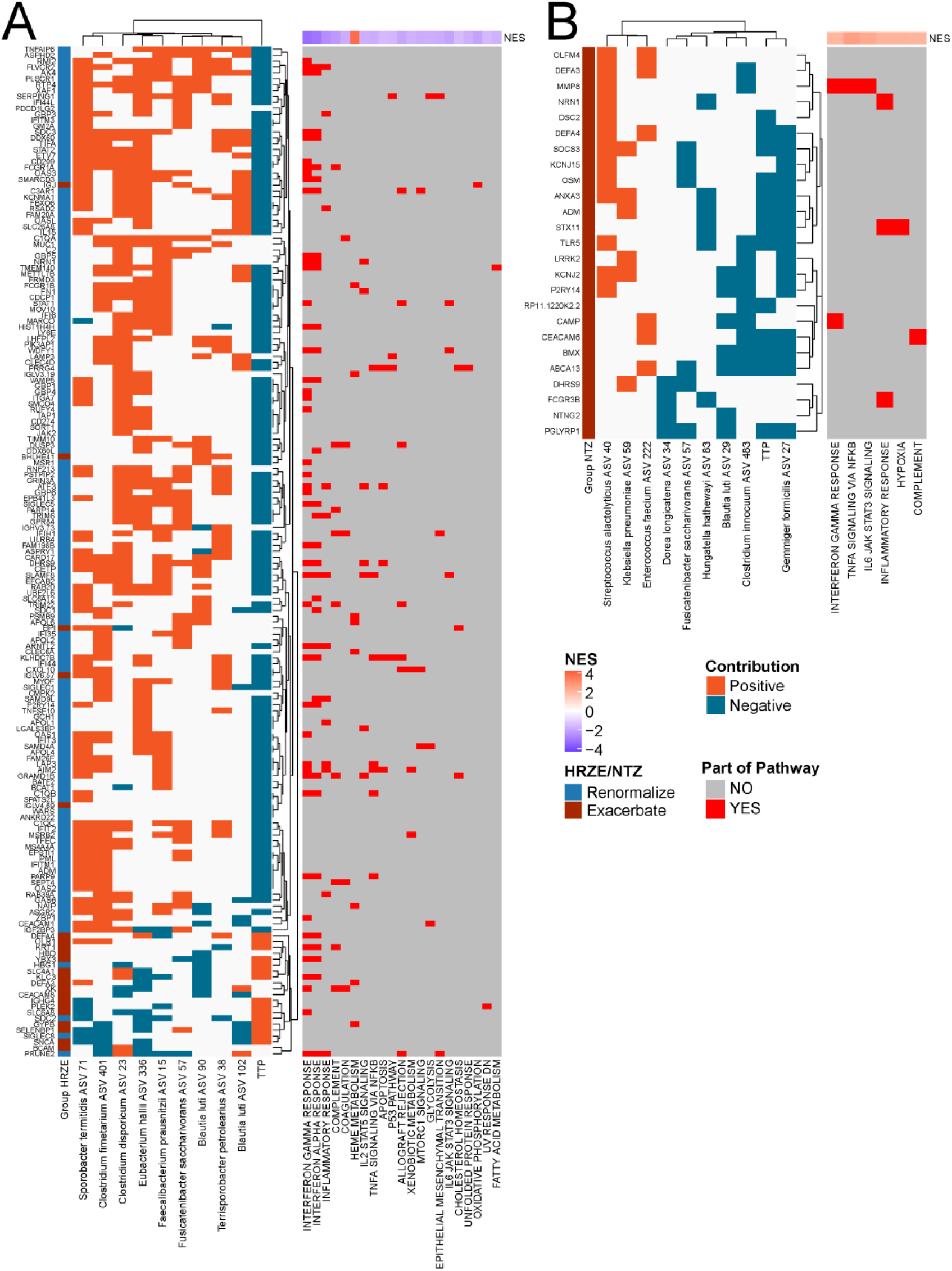
Hallmark Pathway Analysis of peripheral gene transcriptional data and its relationship to gastrointestinal microbiota and *M. tuberculosis* bacterial load (TTP). We used Random Forest regression to model the change in gene expression as a function of gastrointestinal microbiota and reduction in bacterial load (TTP). Top predictors are shown for both HRZE (**A**) and NTZ (**B**) study arms. The contribution of each predictor to a gene’s expression profile is estimated by calculating the derivative (or slope) of the accumulated local effects (ALE) plots between each predictor and each modeled gene (see Methods). The sign of the derivative is displayed in the heatmaps. On the right, the presence or absence of a particular gene in a Hallmark gene set is indicated. If that gene is in the pathway, it is marked as red. Some genes are in multiple pathways, and only the significantly enriched pathways from the differential analysis (of the full transcriptome) are shown. The Normalized Enrichment Score (NES) is how significantly overrepresented a particular pathway is in the treated case (comparing pretreatment vs treatment in both instances) and takes into account pathway size. A majority of the organisms are obligate anaerobic Clostridia, from the following clusters: *Clostridium disporicum* (Cluster I), *Faecalibacterium prausnitzii* (Cluster IV), *Eubacterium hallii* (Cluster XIVa), *Sporobacter termitidis* (Cluster XI), *Fusicatenibacter saccharivorans* (Cluster XIVa), *Clostridium fimetarium* (obligate anaerobe), *Clostridium innocuum* (obligate anaerobe), *Gemmiger formicilis* (obligate anaerobe), *Blautia luti* (Cluster XIVa), *Dorea longicatena* (obligate anaerobe), *Hungatella hathewayi* (obligate anaerobe).

In the model trained on the NTZ treatment data, we find two opposing effects of NTZ induced microbiome perturbation. Depletion of Clostridia by NTZ is predicted to contribute to the exacerbation of inflammatory pathways of active TB observed in the transcriptomic data (Figure 5B: negative contribution in heat map=lower Clostridia>>higher inflammatory transcripts) consistent with the anti-inflammatory properties of these microbiota members. In contrast, the model predicts that pathobionts, whose abundance is enhanced by NTZ (*E. faecium, K. pneumoniae* and *S. alactolyticus*, Figure 2C), have the opposite effect and exacerbate inflammatory transcripts (Figure 5B: positive contribution in heat map=higher pathobionts>>higher inflammatory transcripts). Pathobionts are potentially pathogenic symbionts of the microbiota, and include *E. coli, K. pneumoniae* and *S. alactolyticus*).

Taken together, our data and related computational analyses show that the changes in inflammatory gene expression that accompany treatment of TB may be mediated both by the anti-microbial activity of the drugs that lead to pathogen clearance and by antibiotic induced changes in microbiome composition. Our model identifies two modules of microbiome-inflammatory effects. The first is the exacerbation of TB associated inflammation by depletion of Clostridia, which is evident in both the HRZE and NTZ models. Additionally, the enhancement of pathobionts such as Klebsilla, which only occurs with NTZ, also exacerbates inflammatory pathways. Based on this modelling, we predict that successful disease resolution will be associated with preservation of Clostridia, whereas their depletion and consequent enhancement of Proteobacteria and Bacilli pathobionts might slow resolution or even support inflammatory exacerbation.

### Relationship of the microbiome and peripheral gene expression in a healthy control validation cohort

The results from our primary analysis in the longitudinal treatment cohort demonstrate that specific gastrointestinal bacteria are associated with proinflammatory peripheral blood gene signatures in humans. Specifically, we predict that higher abundance of Clostridia is negatively associated with inflammation (e.g. INFα, INFγ, IL6/JAK/STAT3, Inflammatory Response gene signatures) while high abundance of commonly known pathobionts including *E. coli, Klebsiella, Citrobacter, Streptococcus* and *Enteroccocus* promotes exacerbation of these signatures. To determine the generality of these results we analyzed a control set of human data from two healthy cohorts. A subset of these data were previously reported in our work^5^, and come from a cross-sectional study of healthy household contacts of active pulmonary TB patients (termed Family Contacts, FC) and healthy unexposed donors from the same community in Haiti (termed Community Controls, CC) (see Methods). For these two new cohorts we have a total of 52 healthy control individuals (18 FC and 36 CC) for which we gathered both microbiome 16S rRNA sequencing data as well as peripheral blood transcriptomics for the same individuals.

We analyzed the blood transcriptomics and performed correspondence analysis on the DSEeq2 normalized abundance of the transcripts in each sample (Figure 6A). We observed pretreatment and treated HRZE and NTZ sample clustering away from FC and CC samples, highlighting the fact that individuals with active TB, even while on treatment, represent the transcriptional profile of active TB patients has not returned to a profile of healthy controls (Supplementary Figure S7). More importantly we see that FC and CC samples do not separate in this ordination, suggesting that both cohorts can be used as healthy controls in the subsequent analysis.

**Figure 6:**
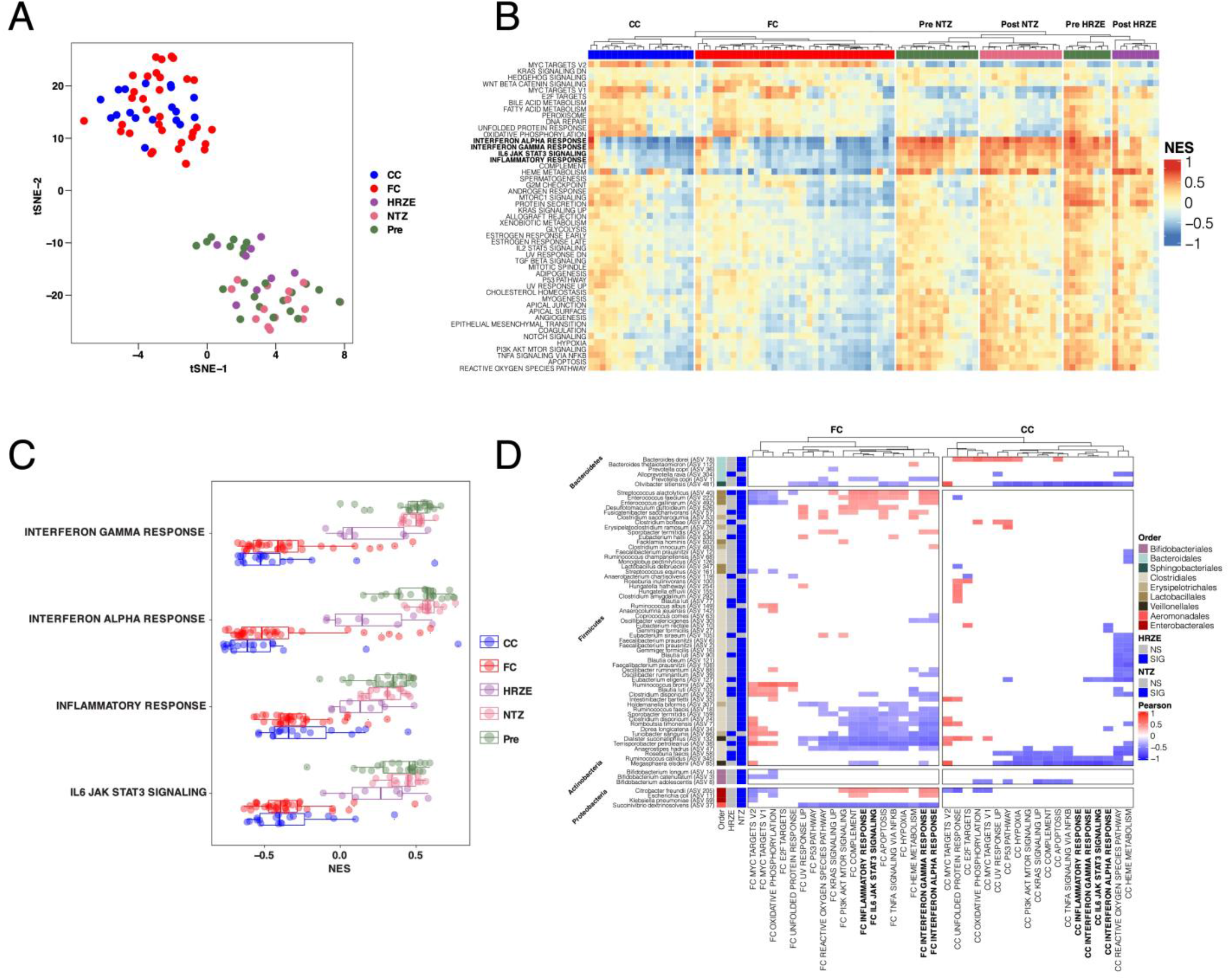
Analysis of microbiome and blood peripheral gene expression in an independent healthy control human cohort validates association between specific microbiome members and host peripheral gene expression. **A**. t-Distributed Stochastic Neighbor Embedding (t-SNE) ordination all RNAseq samples used in this analysis. This highlights the transcriptional differences between active TB patients, even those on HRZE treatment, and the healthy control cohorts. **B**. Within-sample GSEA (ssGSEA) of 50 Hallmark pathways from the MiSigDB on a per-sample basis for all cohorts in this study. NES score was calculated using the variance stabilized transformed counts from DSEeq, and plotted after scaling across all samples. **C**. Four representative pathways that correlated with microbiome composition in all cohorts analyzed in this study. **D**. Pearson’s correlations of ASVs with Hallmark pathways in an unbiased fashion highlighting negative correlation of many Clostridia and the proinflammatory pathways highlighted in 6C.

To link transcript abundance to immune pathway enrichment we utilized gene set variation analysis (GSVA) (see Methods)^35^. GSVA enables computing an enrichment score for any defined list of genes (taken as a surrogate for a biological pathway) in each sample. For each sample, each pathway generates a normalized enrichment score (NES) that can be used for downstream analysis to compare pathway profiling across samples. Importantly, this method is completely agnostic to the phenotype label of the sample.

We performed unsupervised clustering on the samples-by-pathway NES scores, and found that individuals from FC and CC cluster separately from individuals in the longitudinal treatment cohort (Figure 6B). We observed that the NES score signature comparisons between the FC and CC individuals, and individuals before treatment and after two-weeks of HRZE or NTZ have qualitatively reduced enrichment in INFα, INFγ, IL6/JAK/STAT3, and Inflammatory Response (Figure 6B). As discussed, these are the pathways that renormalize with HRZE treatment and exacerbate with NTZ in the longitudinal dataset (Figure 3). These are also the pathways negatively associated with Clostridia abundance and positively with Proteobacteria and Bacilli abundance in the random forest modeling analysis (Figure 5). Interestingly, post HRZE treatment individuals have an enrichment distribution for this pathway which is in between FC and CC individuals and NTZ/Pretreatment samples thus reinforcing the result that HRZE treatment promotes renormalization (Figure 6C).

Even though FC and CC individuals show an overall lower inflammatory enrichment compared to active TB samples, and samples post HRZE and NTZ treatment (Figure 6C), we hypothesize that the FC and CC intragroup inflammatory pathway enrichment distributions would correlate with the abundance of microbes that we found to predict the expression of genes associated with these pathways in the Random Forest Modeling analysis performed on the HRZE and NTZ microbiome/transcriptomic data (Figure 5 and Figure S6). We performed Pearson’s correlation analysis between these NES values, and the abundance of ASVs from bacterial orders having representative species found to be predictive of the abundance of these inflammatory pathways in the longitudinal treatment cohorts (Figure 6D). In these two human cohorts, we again observe a negative association between Clostridia ASVs and INFα, INFγ, IL6/JAK/STAT3, and Inflammatory Response pathways, as well as a positive association between these pathways and the abundance of pathobionts including *Enterococcus, Streptococcus, E. coli* and *Klebsiella*. We interpret these correlations as independent validation of the observation that specific microbiome members are strongly correlated with peripheral inflammatory response pathways in both inflammatory disease states and in homeostatic conditions.

## Discussion

Since the advent of high-throughput microbiome characterization, it has become clear that antibiotics are one of the most common and severe perturbing influences on human microbiome composition, with both acute and longer lasting effects^36,37^. It also has become evident that the specific microbiome constituents have specific effects on host immunity, including the abundance and function of immune cell subsets^21^. Significant prior data have documented the effects of antibiotics on microbiome composition and function and the consequent influence of these microbiome factors on specific immune cell populations^38^, with the majority of these findings derived using *in vivo* mouse models. While there is no doubt that microbiota dynamics affect host immunity^6^, it remains unknown to what degree antibiotic induced perturbation of the microbiome may modify the outcome of treatment of infection. It is conceivable that antibiotics work to clear infection both due to direct pathogen killing and by immune modulation through the microbiome. It is also possible that the pathogen killing effect of antibiotics may be partially counteracted by detrimental immune effects induced by microbiome perturbation. Such dynamics may be particularly relevant to the treatment of chronic infections such at tuberculosis, in which antibiotic therapy is prolonged and the disease manifestations reflect a mixture of pathogen burden and the balance of inflammatory mediators that cause tissue destruction and chronic symptomatology^29,39^.

Antibiotic sensitive tuberculosis is treated with six months of antibiotics with predominantly mycobacterial specific agents. In this study we report the early microbiome effects of HRZE therapy in subjects with active TB and demonstrate that the same changes observed in a human cross-sectional study of TB treatment^19^ (comparing vs cured and LTBI individuals) were present after just two weeks of treatment. As previously shown^19^, we conclude that HRZE treatment has a rapid and narrow effect on the intestinal Class of Clostridia, a finding that was also demonstrated in mice^18,40^. We note that given the mycobacterial-specific nature of TB drugs, and the combinatorial nature in which small molecules interact to affect the microbiome, it was difficult to predict that primarily Clostridia, in the Phylum Firmicutes, would be targeted by HRZE therapy, whereas Actinobacteria, the phylum to which *Mtb* belongs, are relatively unaffected. Experiments in mice have demonstrated that this anti-Clostridia effect is primarily driven by rifampicin^18^. Clostridia are immunologically active components of the microbiota through their production of metabolites such as short chain fatty acids and other compounds^2,5,6,41,42^.

In this work, in addition to determining the microbiome perturbing effect of TB treatment at 2 weeks of therapy, we also leveraged a dataset derived from a clinical trial comparing two TB treatment regimens in a 2-week early bactericidal activity (EBA) format^43^. This comparison allowed us to dissect the relative contributions of pathogen killing and microbiome perturbation to disease resolution because one treatment arm, standard therapy, both reduced *Mtb* bacterial burden and perturbed the microbiome, whereas NTZ had no effect on average *Mtb* burden, but did perturb the microbiome in a fashion that overlapped with HRZE. Additionally, the availability of systemic inflammatory markers derived from peripheral blood transcriptomics allowed us to determine the relative contribution of pathogen sterilization and microbiome disruption in predicting the resolution of inflammatory markers of disease. We find that even though NTZ did not reduce the burden of *Mtb* in the sputum, this molecule nevertheless: 1. caused perturbations responsible for obliterating a large number of obligate anaerobes (e.g. Clostridia), 2. facilitated domination by a number of pathobionts, and 3. affected peripheral gene expression of TB-related and TB-unrelated genes. Specifically, two independently-built computational models (one calibrated on HRZE-treated individuals, the other on NTZ) to link gene expression with microbiota and *Mtb* bacterial load changes showed that changes in active TB transcript patterns were not only correlated with the ability of the drug to reduce *Mtb* bacterial burden, but also with the abundance of Clostridia and pathobionts selected by NTZ. Specifically, the model proposed that reduced *Mtb* load in the sputum and increased abundance of Clostridia are predictive of normalization of the inflammatory transcript profile of active TB. In contrast, increased abundance of pathobionts, including *E. faecium* and *K. pneumoniae*, was predictive of inflammatory exacerbation in the NTZ cohort.

To validate the inferred microbiome-host inflammatory relationship, we mined microbiome and blood transcriptomic profiling from an independent human cohort of healthy Haitian individuals. Remarkably, despite the reduced peripheral levels of inflammatory pathways compared subjects with active TB, we observe that members of the Class Clostridia negatively correlate with pro inflammatory pathways and the reverse for the intestinal pathobionts. This validation data strongly supports our conclusion that microbiome composition sets the tone of systemic inflammation, both in disease states and in homeostatic conditions. Further, it is consistent with the prior findings in both humans and animals that Clostridia have been associated with induction of anti-inflammatory or benign conditions, whereas enrichment in *Enterobacteriaceae* members has been found to be immune-modulating and to alter immune cells populations in the periphery ^44^.

This opposing predictive effects of immune modulating Clostridia and *Mtb* bacterial load (TTP) inferred by the RFR model in HRZE vs. NTZ-treated individuals may be interpreted as follows: when a treatment like HRZE is effectively killing *Mtb*, the rapid reduction in pathogen load in the lung dominates the normalization markers of inflammation, but there are also opposing effects on normalization originating from HRZE-induced microbiome perturbation. In the NTZ group, Clostridia are eradicated, and Proteobacteria and Bacilli are enriched, but without pathogen killing, the microbiota effects are dominant. This NTZ group allows us to deduce opposing effects of Clostridia and pathobionts, with preservation of the former favoring inflammatory resolution and the latter favoring inflammatory exacerbation. This is consistent with our findings that after two weeks of NTZ treatment there is an exacerbation of TB associated inflammatory pathways (Figure 3B).

Finally, given the challenge of explaining the relationship between microbiome composition and peripheral gene expression with paired samples, randomized to drug combinations with vastly different effects on both body systems, we strove to use an appropriate mathematical approach for this type of analysis: random forest regression. While there are a variety of statistical and machine learning techniques able to investigate relationships between complex multiparametric datasets (“large p”: microbiome composition, RNAseq data, clinical metadata, randomization cohort, paired-sample baseline normalization, etc.), and a “small n” of individuals in early phase clinical trials, Random Forests are adequate for microbiome purposes, as they have been shown to outperform Support Vector Machines in some instances, especially for continuous variable data, and need initialization of a smaller set of parameters compared to other deep-learning methods. We believe that our results highlight the utility of these models to: 1. Provide evidence for or against a particular hypothesis about clinically significant relationships between many potentially related parameters, and 2. To provide hypothesis generating relationships between the multi-omic constituents (i.e., features) of these models, which can be further tested in mice, validation cohorts, or other model systems.

Our data indicates that within the first 14 days of treatment of tuberculosis, resolution of the active inflammatory response of TB (as measured by peripheral blood transcriptomics) may be strongly affected both by reducing *Mtb* burden as well as through antimicrobial-induced microbiome perturbations that may act directly on systemic immune function. Among the pathways tightly correlated with both factors are the signature activated pathways of active TB disease: IFNγ, type I interferon, and TNFα ^12^. There is growing evidence that the outcome of active TB reflects a mixture of pathogen burden and cytokine networks that include IL-1 and IFNα, with the latter acting to exacerbate disease^39^. Our findings indicate that the microbiome perturbation that accompanies TB treatment is a predictor of the normalization of these same pathways during early treatment, suggesting that microbiome perturbation could modify or predict the rapidity of disease resolution. In the first two weeks of treatment, pathogen killing is the dominant factor, but microbiome dependent modulation of inflammatory responses during treatment may assume an important role during the later phases of treatment when pathogen killing slows. The validation of the relationships between microbiome composition and peripheral gene expression in a healthy control cohort, especially for the collective expression of these same pro- and anti-inflammatory pathways, suggests that these relationships may extend into other populations. Whether these relationships are causal, or biomarkers of another state will remain at the forefront of future study design. Future work will be directed to applying the analytical tools and study design presented here to later time points in the TB treatment course to examine whether microbiome perturbation during treatment associates with clinically relevant treatment outcomes, and whether the abundance of Clostridia correlates with rapidity of *Mtb* sterilization or the resolution of the inflammatory response that accompanies active TB. Such data might help support trials to test microbiome monitoring as a predictor of TB treatment outcome or help understand interindividual heterogeneity in treatment outcomes.

## Materials and Methods

### Ethical statement and study approval

All volunteers provided written informed consent to participate in this study. All human studies were reviewed and approved by the IRBs of both Weill Cornell Medicine and Groupe Haitien d’etude du Sarcome de Kaposi et des Infections Opportunistes (GHESKIO) Centers (Port-au-Prince, Haiti). Participants provided informed consent prior to peripheral blood draw for whole blood collection and stool collection for 16S rDNA sequencing. All methods and procedures were performed in accordance with the relevant institutional guidelines and regulations.

### Donor recruitment and protection of human subjects

#### Longitudinal treatment cohort

Donors were enrolled through the Clinical Trials Unit at GHESKIO. Pulmonary TB was diagnosed by clinical symptoms, chest radiograph consistent with pulmonary TB, and positive molecular testing. All participant samples were deidentified on site using a barcode system before they were shipped to Weill Cornell Medicine (WCM)/Memorial Sloan Kettering Cancer Center (MSKCC) for analysis. All clinical metadata was collected on site and managed through the REDCap data management system.^45^

#### Human healthy control arm

We recruited families of active pulmonary TB patients where at least 2 siblings within the family were diagnosed with active TB. These criteria were designed to select for households with high risk of transmission of Mtb. Household contacts were then recruited if they had been sleeping in the same house with a TB case for at least 1 month during the 6 months prior to the TB case diagnosis. Contacts underwent clinical screening for active TB symptoms and IGRA testing. Healthy donors without history of TB contacts or disease were recruited from the same community as a control group for exposure and also underwent clinical screening for active TB symptoms and IGRA testing. All donors provided informed consent prior to peripheral blood donation for whole blood collection for RNAseq and stool submission for DNA extraction and 16S rDNA sequencing.

### Microbial DNA extraction from stool

DNA extraction from stool was performed as described.^19^ Stool specimens were collected and stored for less than 24 hours at 4°C, aliquoted (∼2 ml each), frozen at –80°C, and shipped to WCM/MSKCC. About 200 – 500 mg of stool from frozen samples was suspended in 500 μl of extraction buffer (200 mM Tris-HCl [Thermo Fisher Scientific], pH 8.0; 200 mM NaCl [Thermo Fisher Scientific]; 20 mM EDTA [MilliporeSigma]), 210 μl of 20% SDS, 500 μl of phenol/chloroform/isoamyl alcohol (25:24:1; MilliporeSigma), and 500 μl of 0.1-mm–diameter zirconia/silica beads (Biospec Products). Samples were lysed via mechanical disruption with a bead beater (Biospec Products for 2 minutes, followed by 2 extractions with phenol/chloroform/isoamyl alcohol [25:24:1]). DNA was precipitated with ethanol and sodium acetate at –80°C for at least 1 hour, resuspended in 200 μl of nuclease-free water, and further purified with QIAamp DNA Mini Kit (Qiagen) according to the manufacturer’s protocols. DNA was eluted in 200 μl of nuclease-free water and stored at –20°C.

### 16S rDNA sequencing

Primers used to amplify rDNA were: 563F (59-nnnnnnnn-NNNNNNNNNNNN-AYTGGGYDTAAAGN G-39) and 926R (59-nnnnnnnn-NNNNNNNNNNNN-CCGTCAATTYHTTTR AGT-39). Each reaction contained 50 ng of purified DNA, 0.2 mM dNTPs, 1.5 μM MgCl2, 1.25 U Platinum TaqDNA polymerase, 2.5 μl of 10× PCR buffer, and 0.2 μM of each primer. A unique 12-base Golay barcode (Ns) preceded the primers for sample identification after pooling amplicons. One to 8 additional nucleotides were added before the barcode to offset the sequencing of the primers. Cycling conditions were the following: 94°C for 3 minutes, followed by 27 cycles of 94°C for 50 seconds, 51°C for 30 seconds, and 72°C for 1 minute, where the final elongation step was performed at 72°C for 5 minutes. Replicate PCRs were combined and were subsequently purified using the Qiaquick PCR Purification Kit (Qiagen) and Qiagen MinElute PCR Purification Kit. PCR products were quantified and pooled at equimolar amounts before Illumina barcodes and adaptors were ligated on using the Illumina TruSeq Sample Preparation procedure. The completed library was sequenced on an Illumina Miseq platform per the Illumina recommended protocol.

### 16S rDNA bioinformatics analysis

Forward and reverse 16S MiSeq-generated amplicon sequencing reads were dereplicated and sequences were inferred using dada2.^46^ Potentially chimeric sequences were removed using consensus-based methods. Taxonomic assignments were made using BLASTN against the NCBI refseq rna database. These files were imported into R and merged with a metadata file into a single Phyloseq object.

Deposition of data. 16S rDNA sequencing data is deposited with the SRA under accession no. PRJNA445968 (https://www.ncbi.nlm.nih.gov/bioproject/PRJNA445968). Code used for 16S analysis is available at https://wipperman.github.io/TBRU/.

### Computational Analysis

Raw ASV count data was normalized using DESeq2^47^. DESeq2 was used to make differential abundance and expression comparisons between treatment cohorts and between individual pre and post-treatment samples for both microbiota and host gene count data.

To determine how the anti-TB treatment affects both microbiome and peripheral gene expression profiles we performed differential analysis on the counts data obtained by microbiome DNA and peripheral blood RNA sequencing. As the primary endpoint of the clinical trial was powered to determine differences in *Mtb* load (TTP), we had to determine the statistical power available to identify significant differences in the abundance of both microbiota members and in the expression of peripheral genes. We ran power calculations (see Methods) to determine that with 16 microbiome samples and 8 RNAseq samples for the HRZE cohort, with 80% power at α<0.05, we could detect a fold change of 1.4 for microbiome difference and a fold change of 1.8 for mRNA transcripts. In the NTZ cohort, with 18 microbiome samples and 14 RNAseq samples, with 80% power at α<0.05, we can detect a fold change of 1.4 for microbiome differences and a fold change of 1.6 for mRNA transcripts. Additionally, we utilized DESeq in our analyses using baseline normalization within an individual as the gold standard. Power calculations were performed with the RNAseqPower package in R. For microbiome data we calculated a biological coefficient of variation of 0.3, and for RNAseq, we used a coefficient of variation of 0.4. We estimated the expected minimum fold change that we could observe for each group based on the sample size, sequencing depth, and an α < 0.05.

To test whether there were differences between groups, we employed PERMANOVA using the Adonis function in the Vegan R package (https://cran.r-project.org/web/packages/vegan/vegan.pdf), which partitions a distance matrix of ASV count data and runs one-way ANOVA between groups of samples.

To predict the post-pre fold change in abundance of HRZE and NTZ-affected host genes as a function of the corresponding fold change in abundance of microbiota ASVs and TTP using Random Forest Regression.^30^ To perform feature selection for each gene and rank features based on their prediction importance we used Boruta^48^. Boruta is a RF classification and regression wrapper for feature selection that allows identification of variables important for the prediction task while also removing redundant ones. Boruta creates a copy of each independent variable and shuffles them to remove correlation with the original variables (shadow variables). Using this augmented set Boruta builds a RF model and performs a Variable Importance Estimation of all the independent variables (both original and shadow). For every variable it then computes a normalized accuracy score. A true variable is important if its normalized accuracy score is significantly greater than that of shadow variables.

Accumulated Local Effect (ALE) plots were implemented to compute how the change in the expression of each modeled host gene is affected by the level of each predictor identified as important by the forest model. Code to analyze the data and to reproduce all the figures and results is available on Github at https://wipperman.github.io/TBRU/.

### Within sample GSEA analysis

The ssGSEA (single sample gene set enrichment analysis) method^49^ was used to profile within-sample differences between pathways from the MiSigDB Hallmark pathways list^50^ with the GSVA package in R^35^. The MiSigDB Hallmark pathways list is a well validated set of general curated biological pathways that can give insight into specific biological and cellular processes. Additionally, we obtained a list of well validated active TB signatures from the TBSignatureProfilier R package (David Jenkins, Yue Zhao, W. Evan Johnson and Aubrey Odom (2020). TBSignatureProfiler: Profile RNA-Seq Data Using TB Pathway Signatures. R package version 0.0.0.9005. https://github.com/compbiomed/TBSignatureProfiler. Variance stabilized transformed (vst) counts derived from DESeq2 were used as input into the GSVA function in the GSVA R package with default parameters and scaled Normalized Enrichment Scores (NES) were plotted as heatmaps. Importantly, unlike classical GSEA, this analysis is agnostic to sample phenotype.

## Data Availability

Deposition of data. 16S rDNA sequencing data is deposited with the SRA under accession no. PRJNA445968 (https://www.ncbi.nlm.nih.gov/bioproject/PRJNA445968)
Code used for 16S analysis is available at https://wipperman.github.io/TBRU/.
Code to analyze the data and to reproduce all the figures and results is available on Github at https://wipperman.github.io/TBRU/.

## Author Contributions

Patient recruitment, enrollment, and sample collection were contributed by LM, KM, KFW, JB, and SCV; Laboratory experiments were performed by MFW and CKV; Data analysis was performed by MFW, SB, VB; Wrote manuscript: MFW, SB, MSG, VB; Edited manuscript: all authors.

MFW and SB are co-first authors, and MSG and VB are co-last authors. Co-authorship and author order were determined by recognition that the integration of the nuances clinical trial data and mathematical modeling are different skillsets found in different laboratory environments. Each were important components to the validity and message of this manuscript.

## Supplementary Figures

**Supplementary Figure S1:**
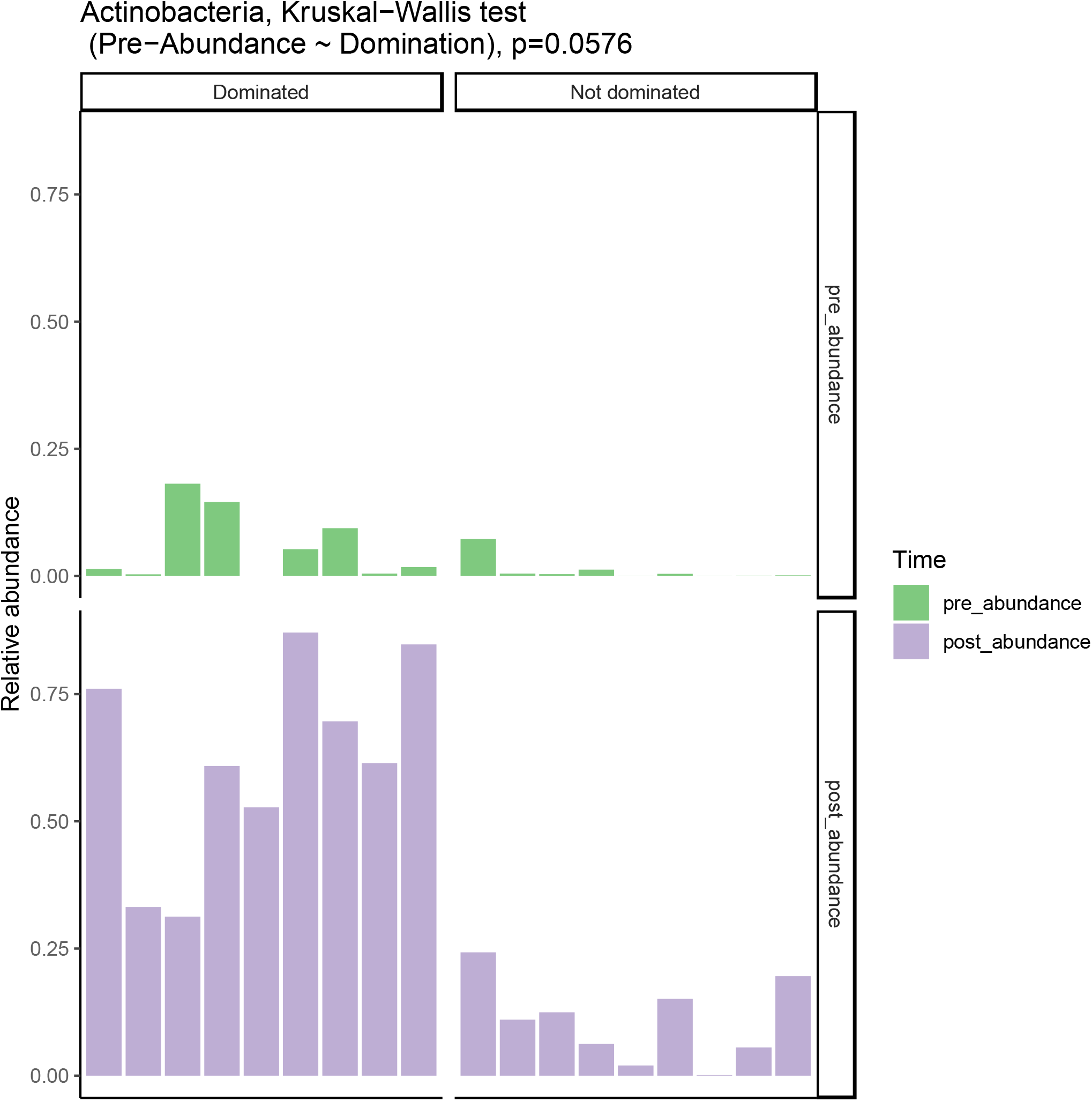
Pretreatment abundance of Phylum Actinobacteria predicts post-treatment domination status (p=0.0576).

**Supplementary Figure S2:**
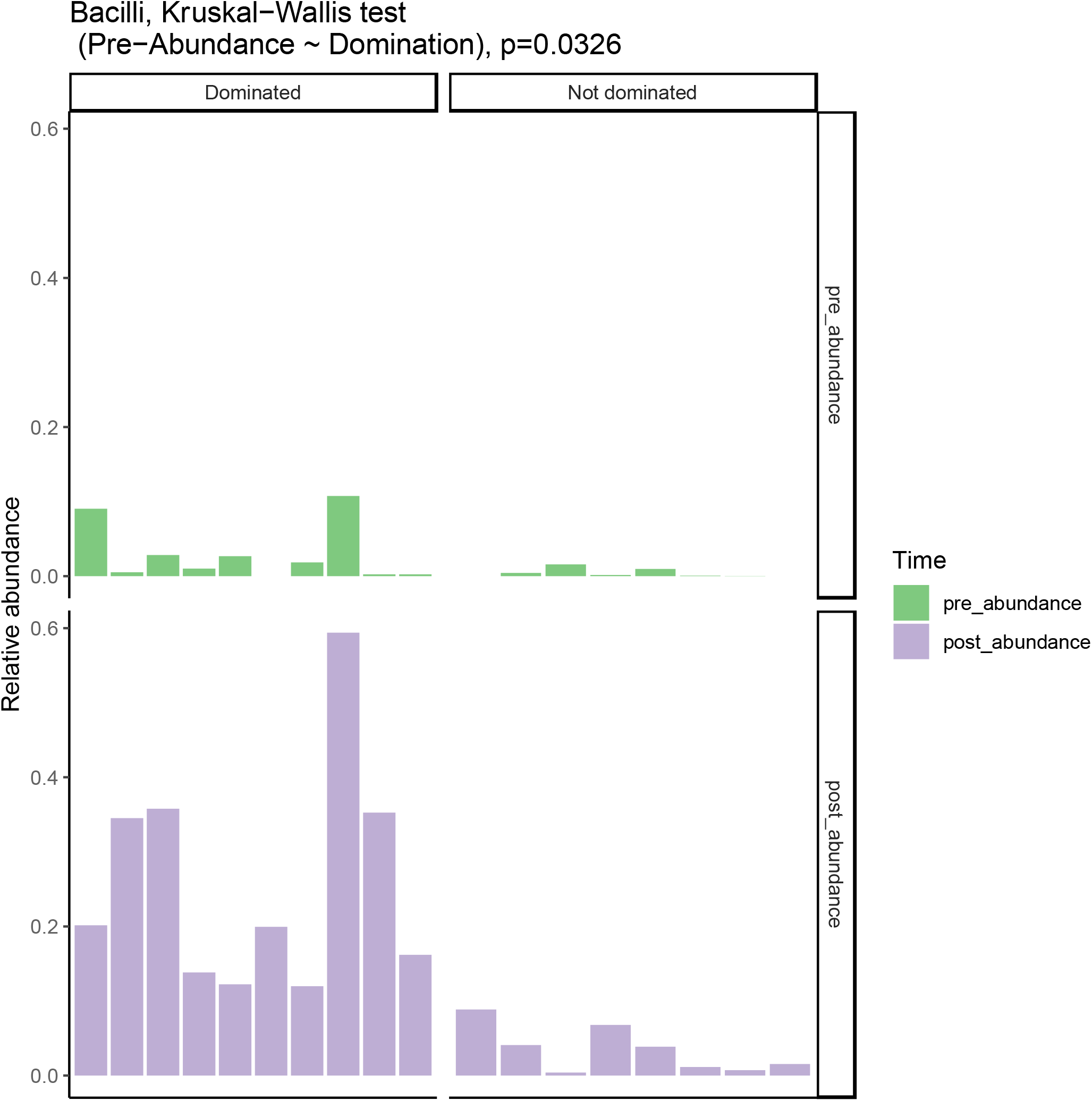
Pretreatment abundance of Class Bacilli predicts post-treatment domination status (p=0.0326).

**Supplementary Figure S3.**
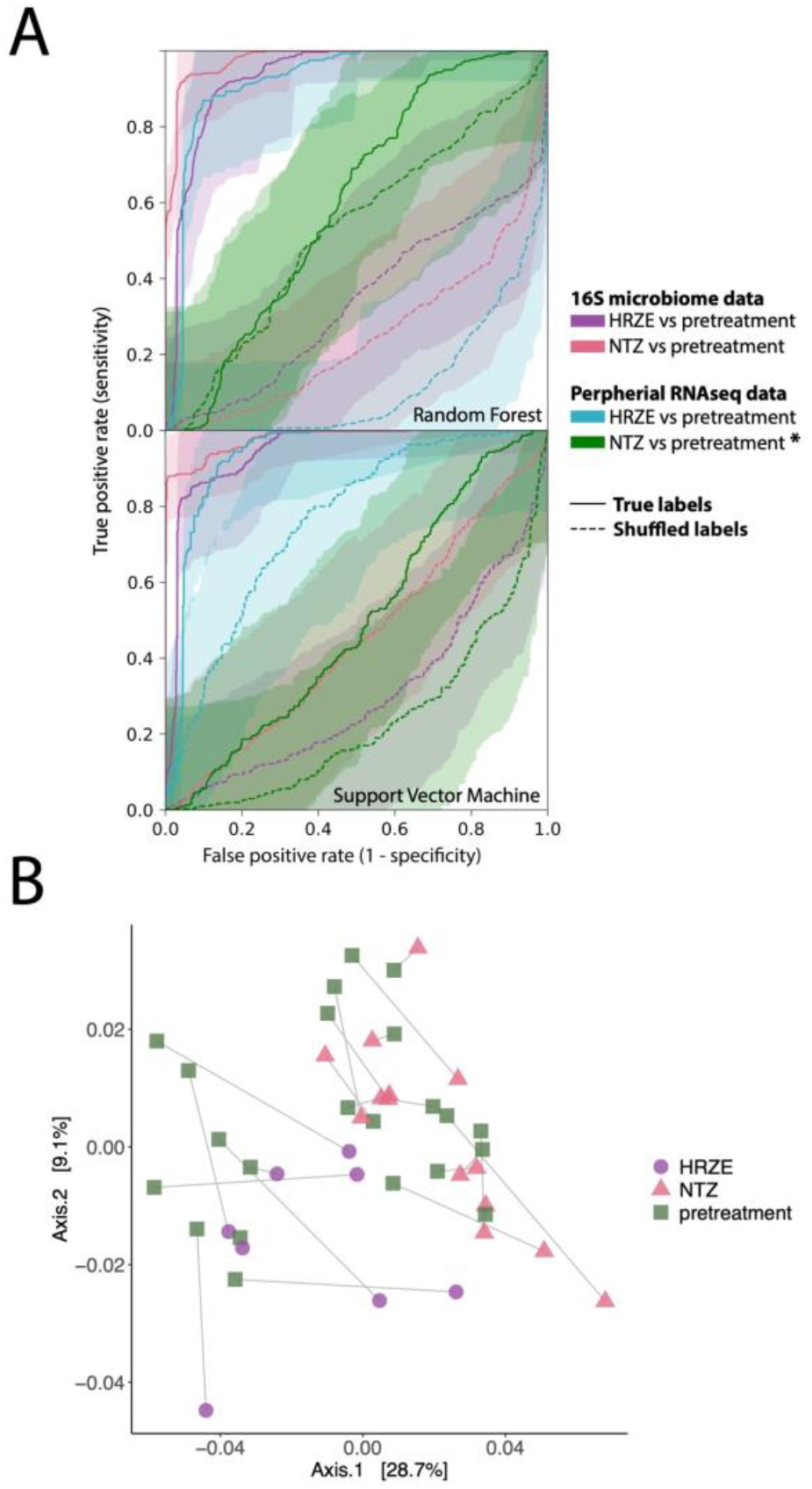
**A**. Receiver operating characteristic (ROC) curves with confidence intervals for random forests and support vector machines generated through comparison of microbiome data and peripheral RNAseq gene expression data for volunteers before treatment and after 14 days of either HRZE or NTZ therapy. Solid lines indicate the true comparison, and dotted lines indicate random (scrambled) labels. **B**. PCA plot of RNAseq peripheral gene expression comparing pretreatment samples randomized to either NTZ or HRZE.

**Supplementary Figure S4.**
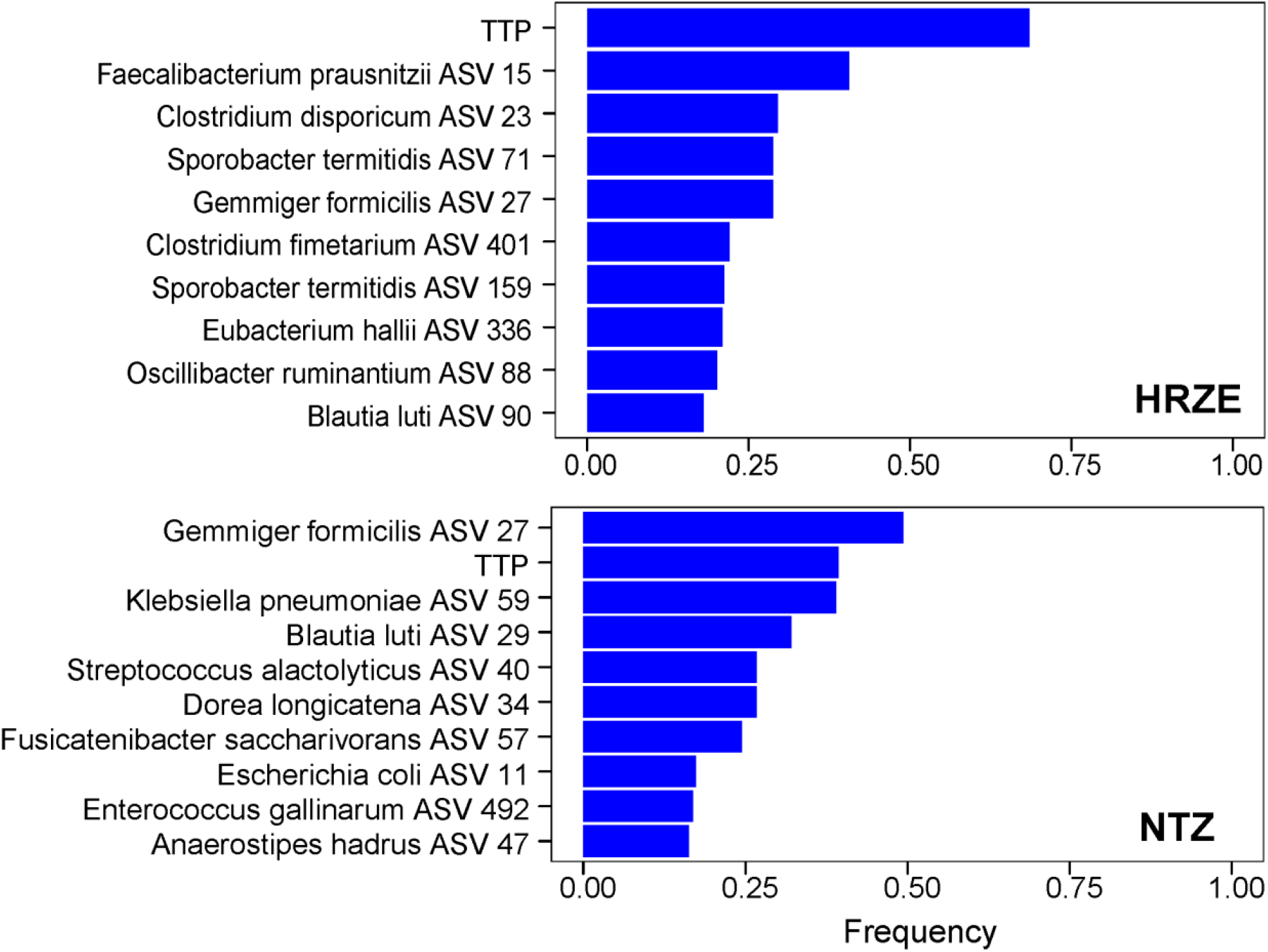
Summary of Random Forest Regression results for all differentially expressed transcripts between pretreatment/posttreatment samples (not just those previously associated with active TB) that were found significantly affected by HRZE or NTZ (See Methods). Frequency indicates the ratio between the number of times a predictor is found to be significantly important in predicting gene abundance over the total number of surveyed genes.

**Supplementary Figure S5.**
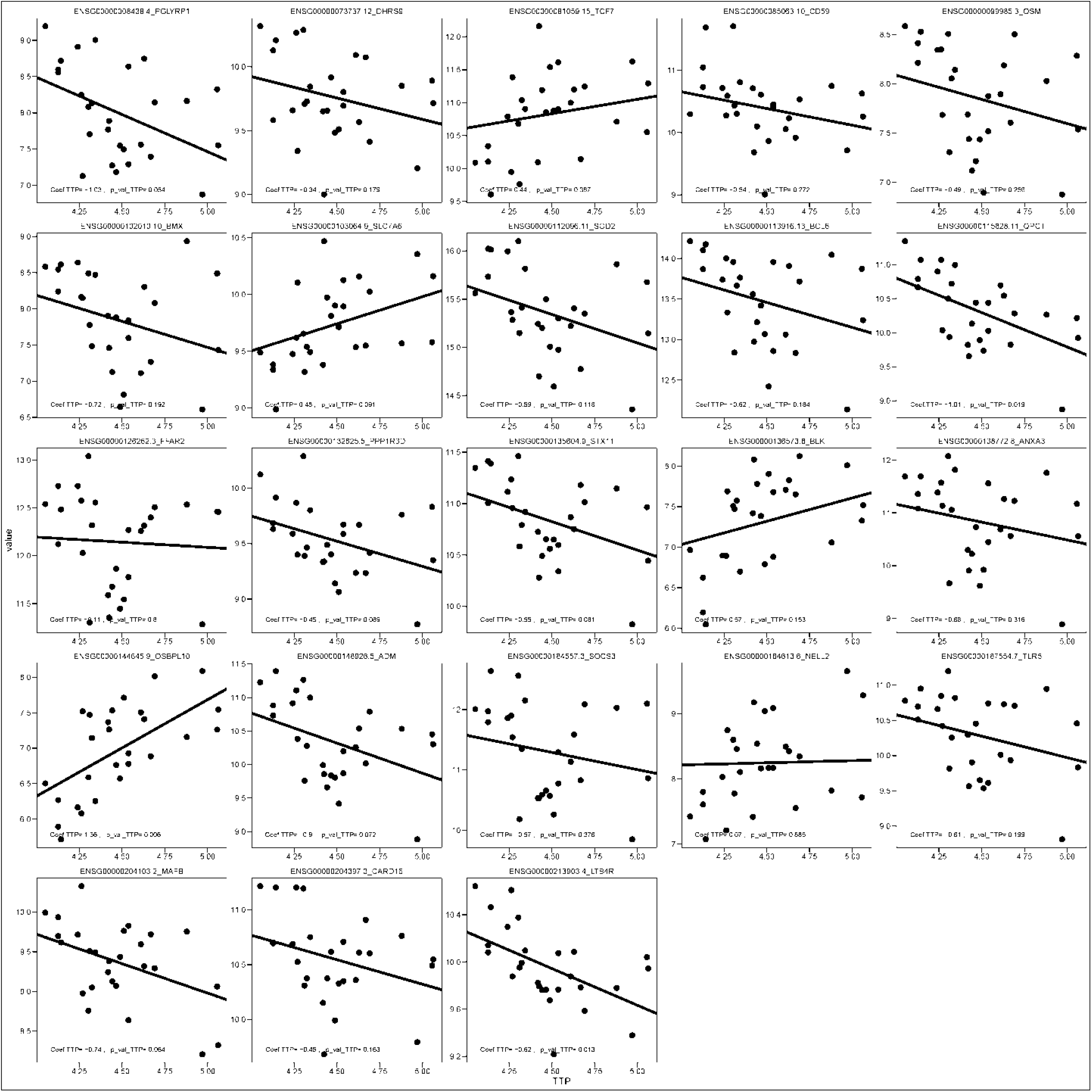
Application of linear mixed effect modeling to predict the abundance of transcripts that are found to be significantly affected by TB bacterial load as measured by TTP in the NTZ cohort as a function of NTZ and by using patient ID as random effect. Even though NTZ does not affect TTP, variability in change in TTP for these individuals is found to explain changes in expression for several host genes previously associated with active TB. The y axis shows the change in each gene’s expression from 0 to 14 days, and the x axis shows the change in TTP across that same time.

**Supplementary Figure S6:**
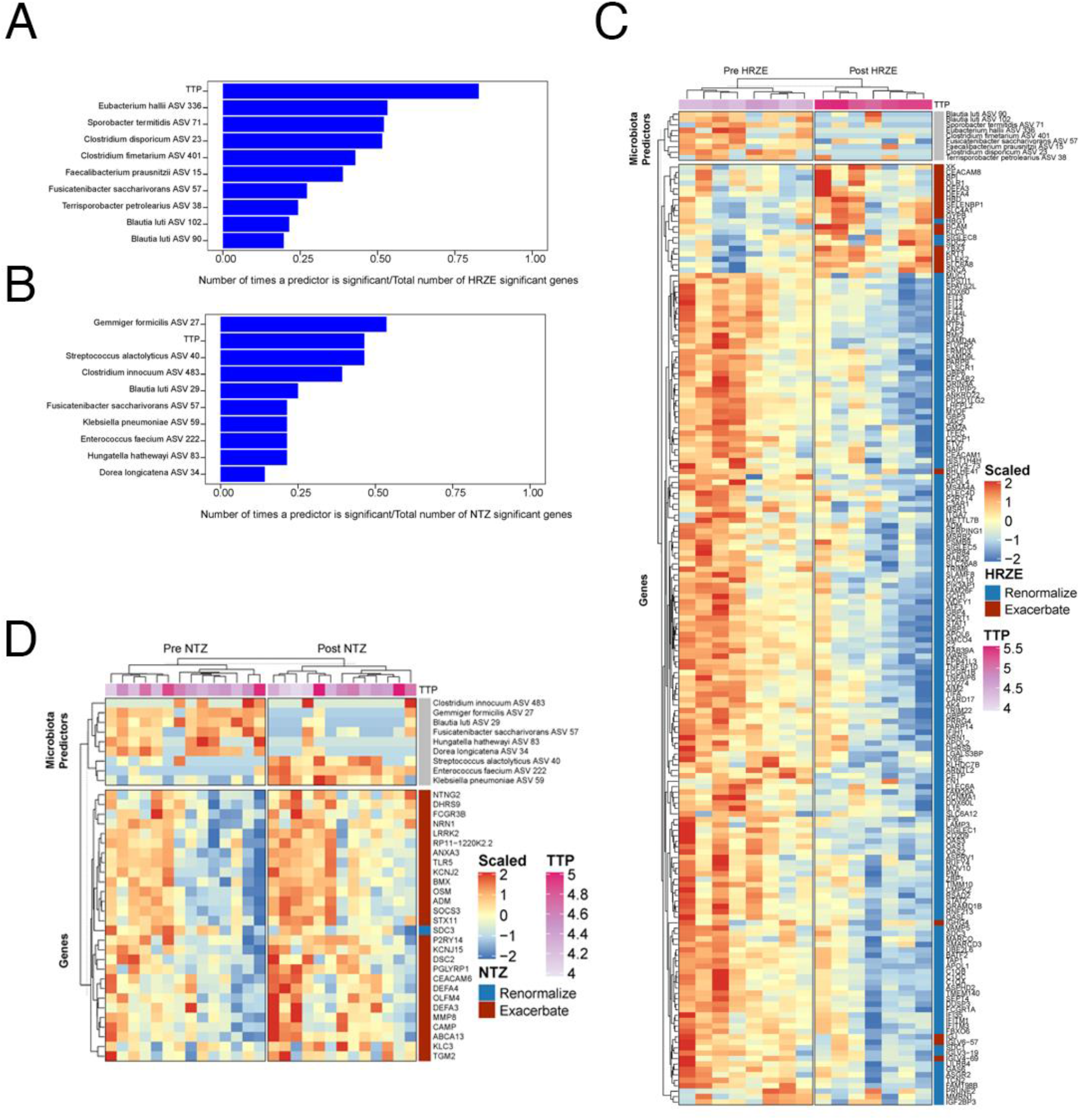
Peripheral gene expression changes in HRZE and NTZ treated groups. **A**. Top 10 predictors of gene expression change in the HRZE cohort. Bacterial load (TTP) is the primary quantity that predicts the change in gene expression for almost three quarters of the genes that change in the HRZE cohort, with significant contributions of Clostridia. **B**. Top 10 predictors of gene expression change in the NTZ cohort. Change in abundance of *Gemmiger formicilis* is the primary quantity that predicts change in gene expression in the NTZ cohort. **C**. Scaled relative abundance of the top predictors and the genes that are significantly altered for HRZE. **D**. Scaled relative abundance of the top predictors and the genes that are significantly altered for NTZ.

**Supplementary Figure S7:**
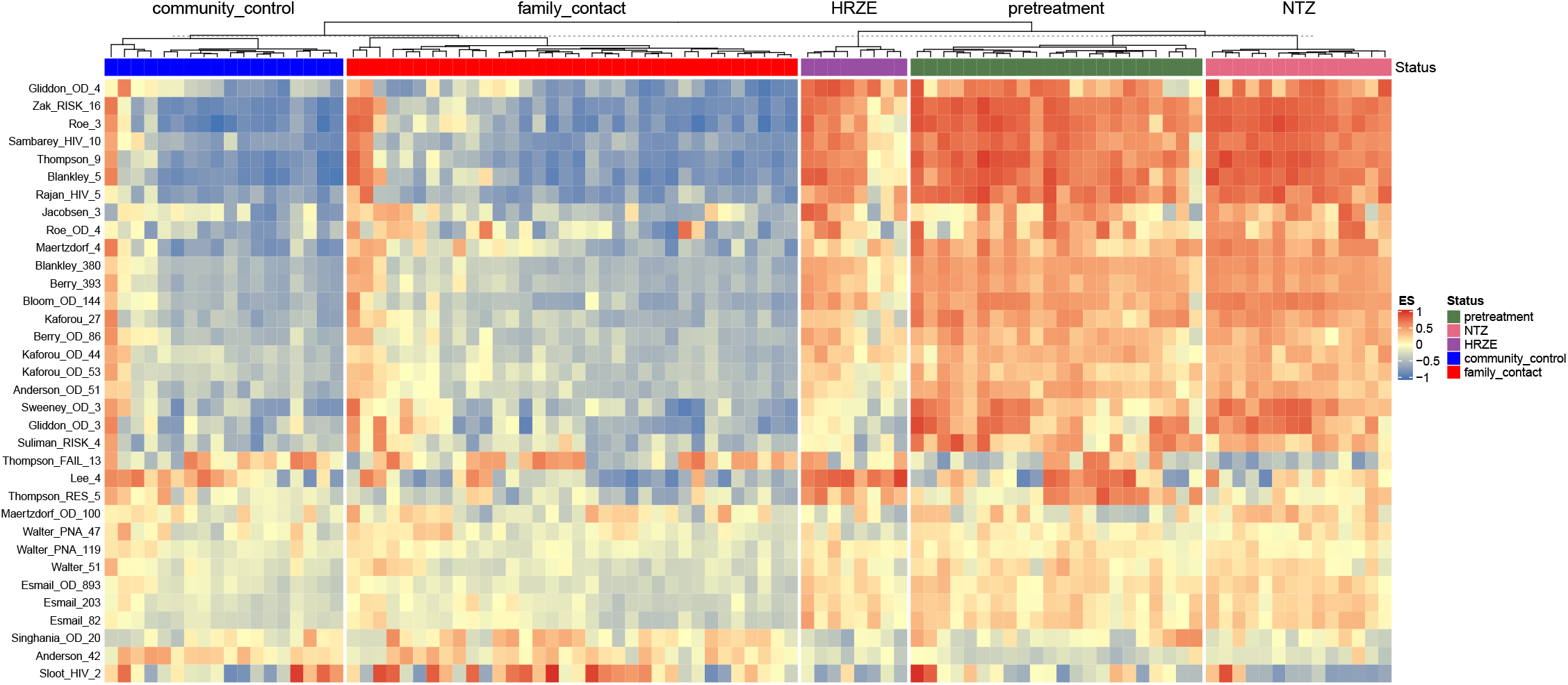
Within-sample GSEA analysis (ssGSEA) of common TB pathways from the R package TBSignatureProfiler. These pathways represent well validated gene lists that predict active TB from LTBI progression, and were the inputs used to generate the meta-signature in the primary analysis of this paper ^17^.

